# Long-Term Effects of Chronic Hemiparetic Stroke and Botulinum Neurotoxin on Wrist and Finger Passive Mechanical Properties

**DOI:** 10.1101/19011312

**Authors:** Benjamin I Binder-Markey, Wendy M Murray, Julius P.A. Dewald

## Abstract

**Background:** Neural impairments that follow hemiparetic stroke may negatively affect passive muscle properties, further limiting recovery. However, factors such as hypertonia, spasticity, and botulinum neurotoxin (BoNT), a common clinical intervention, confound our understanding of muscle properties in chronic stroke.

**Objective:** To determine if muscle passive biomechanical properties are different following prolonged, stroke-induced, altered muscle activation and disuse.

**Methods:** Torques about the metacarpophalangeal and wrist joints were measured in different joint postures in both limbs of participants with hemiparetic stroke. First, we evaluated 27 participants with no history of BoNT; hand impairments ranged from mild to severe.

Subsequently, seven participants with a history of BoNT injections were evaluated. To mitigate muscle hypertonia, torques were quantified after an extensive stretching protocol and under conditions that encouraged participants to sleep. EMGs were monitored throughout data collection.

**Results:** Among participants who never received BoNT, no significant differences in passive torques between limbs were observed. Among participants who previously received BoNT injections, passive flexion torques about their paretic wrist and finger joints were larger than their nonparetic limb (average interlimb differences = +42.0±7.6SEM Ncm, +26.9±3.9SEM Ncm, respectively), and the range of motion for passive finger extension was significantly smaller (average interlimb difference = -36.3°±4.5°SEM; degrees).

**Conclusion:** Our results suggest that neural impairments that follow chronic, hemiparetic stroke do not lead to passive mechanical changes within the wrist and finger muscles. Rather, consistent with animal studies, the data points to potential adverse effects of BoNT on passive muscle properties post-stroke, which warrant further consideration.

## Introduction

Following a hemiparetic stroke, induced damage to corticofugal motor (i.e., corticospinal and corticobulbar) pathways result in an increased reliance on indirect contralesional corticoreticulospinal pathways.^1-5^ This shift alters neural input to spinal motor neurons resulting in motor impairments such as weakness,^6-8^ loss of independent joint control,^9-13^ and motor neuron hyperactivity manifesting as hypertonia and spasticity (i.e., hyperactive stretch reflexes).^14-18^ This altered motor drive and associated limb use, over time, may change muscle structure and function, further amplifying the brain injury induced motor impairments.

Additionally, botulinum neurotoxin (BoNT) chemical denervation, widely used to treat muscle hypertonia and spasticity,^19-21^ may also alter muscle.^22-29^ One particularly detrimental sequelae of these muscle adaptations are that muscles can become increasingly stiff, making movement progressively more difficult.

There is not a clear consensus regarding how prolonged exposure to altered neural inputs following a stroke affect muscle structure and its associated passive mechanical properties. Previous studies quantifying muscle adaptions following a stroke have demonstrated decreased fascicle lengths,^30-32^ often accompanied by increased joint torques and stiffness^7,30,31,33-37^. Such data has been interpreted as evidence of muscle contractures and mechanical property adaptations. However, conflicting evidence also suggests that passive mechanical properties are not different between the paretic and non-paretic limbs of stroke survivors.^33,38^

Abnormal muscle hypertonia and spasticity increase the chance of muscle activity, even in conditions intended to be “passive” or “relaxed”. Thus, conflicting results could be a consequence of differences in how effectively abnormal muscle hyperactivity was controlled among different studies. For example, some studies utilized continuous motion to collect passive torques.^7,35,36^ However, even low constant velocities elicit hyperactive stretch reflexes that substantially increased paretic finger torques.^8^ In other studies, EMGs were not recorded to confirm muscles were passive during data collection.^30,38^ Additionally, some methods involved dynamic analytical models to decompose active torques and estimate passive torques from muscle activity and EMGs estimated rather than directly measuring passive torques.^7,34,36^ Finally, while pre-stretching attenuates hyperactive motoneurons and the stretch reflex,^39^ few studies discuss whether stretching prior to data collection was utilized.^7,30,35,36,38^ Such methodological differences among previous studies obfuscate our understanding of muscle passive properties following chronic stroke.

An additional confounding factor, neither discussed nor reported in the context of the inclusion/exclusion criteria in previous studies, is the potential long term effects of BoNT. BoNT chemical denervation is included in neurological clinical practice guidelines for treatment of adult spasticity^19^; it is a common treatment for muscle hypertonia and spasticity^19-21^ within the ∼45% of stroke survivors who have spasticity.^40,41^ Presumably, the short term, BoNT-induced reduction of muscle tone^20^ improves function by increasing range of motion (ROM) and reducing overall hypertonicity. Yet, evidence of these beneficial effects is limited and based largely on acute, subjective clinical assessments of muscle tone and ROM without demonstrating improvements in either active function or quality of life.^21,42,43^ In contrast, both acute and chronic structural changes in rodent muscles have been observed following BoNT injections, including increased intramuscular connective tissue and passive mechanical stiffness.^22-26,28,29^ If BoNT administration imposes the same effects within human muscle, it is critical to understand both how these structural changes are distinct from stroke-induced alterations and the extent to which the induced structural changes may effect optimal functional recovery.

The overall objective of this study is to determine the extent to which muscle passive biomechanical properties in the paretic limb differ from non-paretic limb given prolonged, stroke-induced, altered muscle activation and use; as well as potential long-terms effects of BoNT administration. To do so, we compared the passive torques collected about the paretic and non-paretic wrist and finger joints of individuals with chronic hemiparetic stroke, while controlling for muscle hyperactivity and accounting for BoNT use. We chose to study wrist and finger muscles because they are affected by the longest lasting and most severe motor impairments following a stroke^44-47^ and are a frequent site of BoNT treatment. To evaluate the long term effects of stroke-induced neural impairments, we recruited individuals with mild to severe hand impairments with no history of BoNT injections. To understand long-term effects of BoNT, and resulting chemical denervation, we recruited additional chronic stroke participants with a history of BoNT treatment. Based on the prevailing findings of previous literature and the common clinical presentation of stiff, flexed wrists and fingers in the paretic limb following stroke, our primary hypothesis is that passive torques in the paretic hand are greater than the non-paretic hand.

## Materials and Methods

### Participants

The Clinical Neuroscience Research Registry from Northwestern University and the Shirley Ryan Ability Lab was used to pre-screen individuals with chronic hemiparetic stroke based on set inclusion/exclusion criteria (Table 1). 105 individuals whose registry-based, prior clinical assessments of upper limb impairment fit the screening criteria were contacted; 68 individuals responded. In this screening (no-BoNT), previous history of BoNT injections in their arms was an exclusion criteria. After registry-based stratification into different impairment levels (mild, moderate, and severe) and further verification that inclusion/exclusion criteria were met 39 individuals were invited; 28 agreed to participate and were enrolled. Once enrollment of the no-BoNT participants was complete, we contacted 23 individuals with a previous history of BoNT injections. Eleven individuals met our enrollment criteria for the BoNT-injected group; 10 were enrolled. Each of these participants received BoNT injections within their forearm flexor compartment. A single enrollee in the BoNT group was subsequently excluded due to also having received BoNT injections in the forearm extensor compartment. The 9 remaining individuals were at least one year post-BoNT injection. After testing was completed on all enrolled participants, the data from three additional enrollees (2, BoNT-injected; 1, no-BoNT) were excluded from the analysis due to the inability to relax during testing, as determined by EMG activity.

**Table 1:**
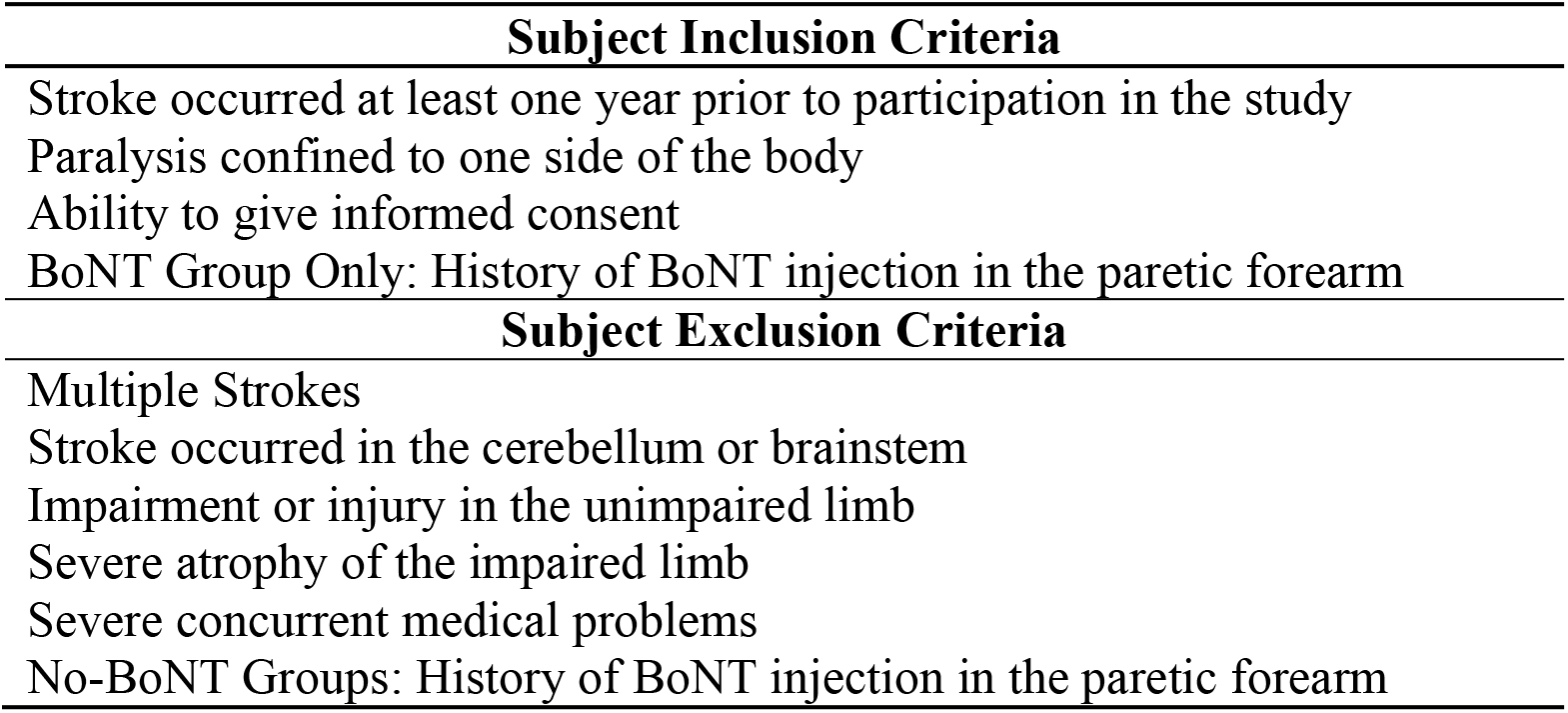
Table of Inclusion/Exclusion Criteria for Participants.

| **Subject Inclusion Criteria** |
| --- |
| Stroke occurred at least one year prior to participation in the study |
| Paralysis confined to one side of the body |
| Ability to give informed consent |
| BoNT Group Only: History of BoNT injection in the paretic forearm |
| **Subject Exclusion Criteria** |
| Multiple Strokes |
| Stroke occurred in the cerebellum or brainstem |
| Impairment or injury in the unimpaired limb |
| Severe atrophy of the impaired limb |
| Severe concurrent medical problems |
| No-BoNT Groups: History of BoNT injection in the paretic forearm |

Enrolled study participants were ultimately stratified by hand impairment using the Chedoke McMaster Stroke Assessment Hand Score (CMSA-HS)^48^, administered at the time of testing by a licensed physical therapist (Table 2). Severe impairments were defined as CMSA-HS scores of 1-3 (n=9 no-BoNT; n=7 BoNT-injected), moderate impairments as 4-5 (n=9 no-BoNT), and mild impairments as 6-7 (n=9 no-BoNT). Clinical scores (both CMSA-HS and Modified Ashworth Scale (MAS)) for all 7 participants with a history BoNT injections indicated severe impairment; scores were not significantly different (p=0.196, p=0.158, respectively) from those with severe impairments and no history of BoNT (Table 2). Demographic data were gathered from all participants prior to data collection.

**Table 2:** Subject Demographics. (CMHS – Chedoke McMaster Stroke Assessment Hand Score, MAS – Modified Ashworth Scale)

| Impairment Level | Sex | Age in years (SD) | Time Since Stroke in years (SD) | Age at time of stroke in years (SD) | Paretic Side | CMHS | MAS | Time Since Last BoNT Injection in years (SD) |
| --- | --- | --- | --- | --- | --- | --- | --- | --- |
| BoNT-Injected Severe (n=7) | 6-M, 1-F | 56.7 (8.3) | 10.7 (5.3) | 46.0 (5.1) | 4-L,3-R | 2.85 (0.38) | 2.71 (0.48) | 4.5 (2.8) |
| No-BoNT Severe (n=9) | 6-M, 3-F | 60.28 (10.40) | 17.1 (7.91) | 43.2 (13.6) | 5-L, 4-R | 2.44 (0.73) | 2.17 (0.87) | n/a |
| Moderate (n=9) | 5-M, 4-F | 64.3 (7.95) | 13.2 (7.96) | 51.1 (12.5) | 6-L, 3-R | 4.22 (0.44) | 1.78 (0.94) | n/a |
| Mild (n=9) | 5-M, 4-F | 56.93 (12.18) | 8.41 (3.71) | 48.5 (9.8) | 1-L, 8-R | 6.44 (0.53) | 0.28 (0.57) | n/a |

Initial enrollment targets (n = 9 per group: no-BoNT mild, moderate, severe, and BoNT-injected) were defined to surpass the conditions needed to detect a large effect size (Cohen’s d = 1.1), based on a priori power analyses of a one-tailed, within-subject t-test for the hypothesis that passive torques in the paretic limb are greater than the non-paretic limb. Due to the lack of appropriate passive torque data, we powered the study based on the detectable interlimb differences in the passive range of motion limits for MCP extension. Specifically, Cohen’s d was calculated from the angular resolution of the device used to measure MCP torque (15°) and intersubject variability at the limit of MCP extension in nonimpaired participants.^49^ The analysis indicated that with our large, standardized effect size, a power of 0.8 would be achieved with α = 0.05 and 7 participants.^50^ With the same assumptions, an a priori power analysis for one-way (between groups) ANOVA indicated that a total of 16 participants were needed.^50^ The total participant numbers for the results presented here (n = 34) are more than double the total specified via the a priori ANOVA analysis. Participant numbers within each of the 4 distinct groups either meet or exceed the number specified via the a priori analysis for a single t-test (n = 7).

The study protocol was developed in compliance with the Declaration of Helsinki and approved by the Institutional Review Board (IRB) of Northwestern University (IRB Study: STU00203691). Participants gave informed consent prior to participation and all patients signed a Patient Consent-to-Disclose form prior to any images being taken.

### Experimental Set-up

A custom built device^51^ was used to collect torques produced about the wrist and fingers. The device allows the experimenter to position the wrist and metacarpophalangeal (MCP) joints separately, in discrete 15° increments, while simultaneously collecting torques about each joint. Participants were seated in an upright position with their hand secured in the device. The participant’s arm was positioned comfortably at their side with the forearm parallel to the ground and palm facing medially. The two distal finger joints were splinted. Muscle activity was monitored throughout each trial using surface electrodes (16-channel Bagnoli EMG System, Delsys Inc., Boston, MA; 1000 x gain, 20-450 Hz bandpass) placed over 4 muscles; Flexor Digitorium Superficialis, Flexor Carpi Ulnaris, Extensor Digitorium Communis, and Extensor Carpi Radialis Longus.

### Muscle Hyperactivity Inhibition Protocol

Muscle hyperactivity was reduced and quieted during data collection by providing conditions that encouraged participants to sleep. This relaxed state reduces reticulospinal tract activity resulting in decreased spinal motoneurons excitability and muscle hyperactivity.^52-54^ We encouraged sleep by creating a dark atmosphere and playing relaxing videos or music. Each session began with 10-15 minutes of stretching of the shoulder, elbow, wrist, and fingers muscles. Participants were then positioned into the device. The lights were turned off, the video or music was turned on, and these conditions remained in effect for the remainder of the protocol. An additional 10 minutes of 90 second sustained stretches of the wrist and fingers muscles were performed, accomodating the individual to the device. Decreases in EMG activity and measured torques were observed at this point (Figure 1). The device’s MCP and wrist restraints were unlocked, allowing the wrist and fingers to move and rest at their equilibrium postures. Three 15-second baseline trials with no evidence of EMG activity were then collected to zero the device and define passive EMG baseline.

**Figure 1:**
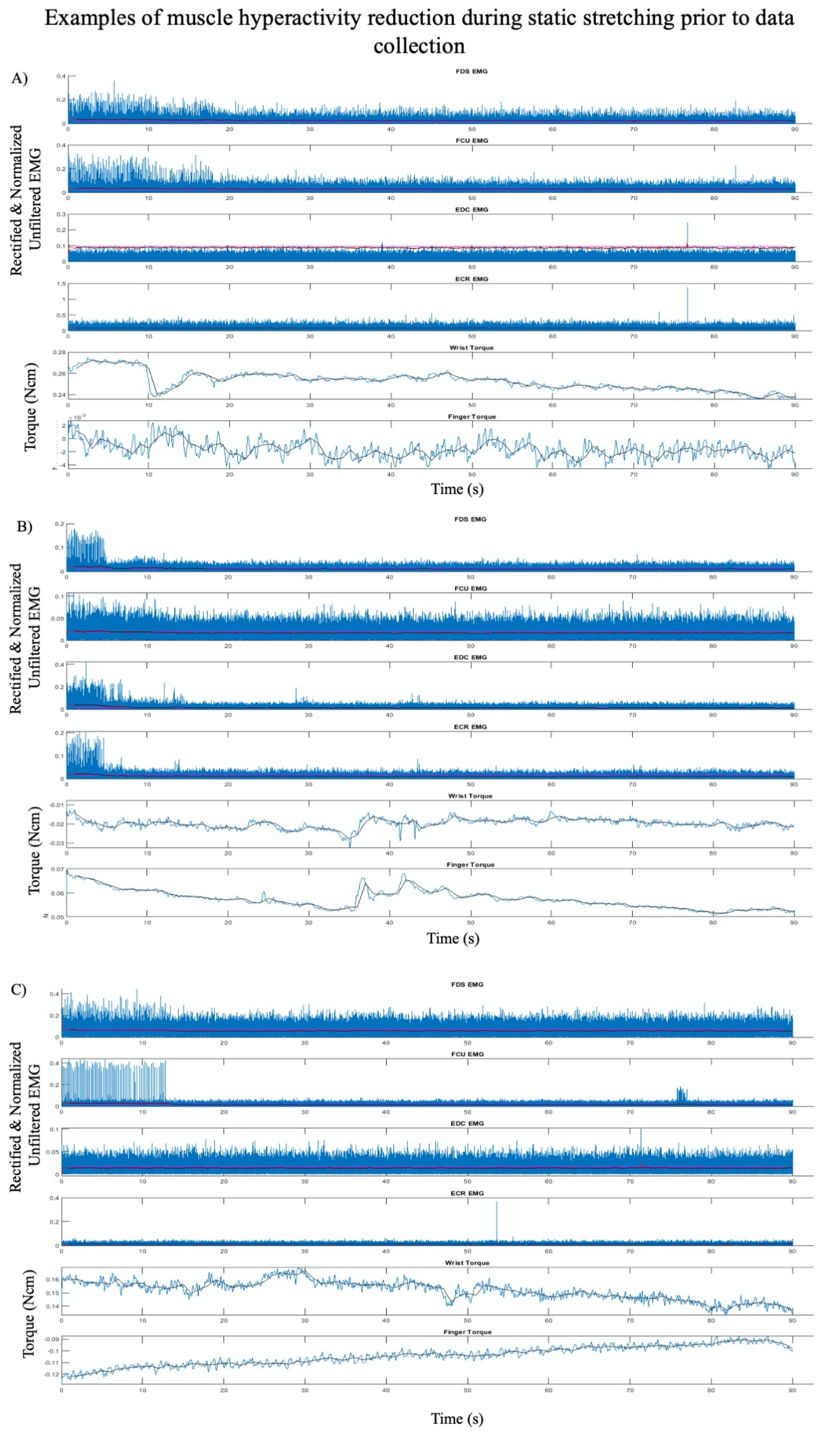
Representative plots, from three separate individuals (A, B, & C), of EMG and torque traces over 90 seconds within one of the initial stretches accommodating the participant to the device and reducing muscle hypertonia. Displayed are raw rectified unfiltered data (blue lines), rectified and filtered EMG (red lines), processed 1 second binned data (black lines), with EMG threshold cutoff for active muscle (magenta lines). Though the participant was “relaxed” at the beginning of each stretch there is significant EMG activity initially with subsequent EMG and torque reduction within each. Example A demonstrates Flexor Digitorium Superficialis (FDS) and Flexor Carpi Ulnaris (FCU) EMG initially firing at a low levels with activity decreasing and a corresponding decrease in wrist torque that continues throughout the trial. Example B demonstrates EMG activity reducing across all muscles with a corresponding decrease in finger and wrist torque that continues throughout the trial. Example C demonstrates low levels of FDS and high levels of FCU EMG hyperactivity initially with hyperactivity stopping within 15 seconds and a corresponding decrease in wrist extension torque that continues throughout the trial.

### Experimental Procedure

The protocol for each limb was completed on two consecutive days, if possible. Data collection took four to six hours per limb. Data from the non-paretic hand was collected on the first day to acclimate the individual to the procedure. Because of the sensitive nature of muscle hyperactivity in the paretic limb, we felt the benefits of testing that limb after the participant was fully comfortable with the full protocol and able to maximally relax outweighed any costs associated with not randomizing for limb across days. Passive torques were collected from a maximum of 108 combinations of wrist and MCP joint postures, depending on the individual participant’s available ROM. MCP and wrist ROM were determined within the device; we recorded the largest 15° increment reached for each degree of freedom. The most extreme postures that could be reached by each participant within the device were determined by the individual’s comfort level or the device’s limits.

Torques were collected at static postures. Wrist posture was randomly set in 15° increments between 60° of flexion and 60° of extension (9 wrist postures). One participant (BoNT-injected) had limited wrist extension in the paretic limb, and could only be positioned in 6 postures. At each wrist posture, the MCP joints were positioned in full extension, moved to full flexion, and then moved back to full extension in static 15° increments. At each static posture, data were collected for 15 seconds and visually inspected. Trials for which there was clear evidence of muscle activity and corresponding torque deviations during testing were discarded and repeated.

### Data processing

Raw torque and EMGs were collected and digitized (CED Micro 1401 MkII, Cambridge Electronic Design, Cambridge, UK) at a 1 kHz sampling frequency using Spike2 software (Cambridge Electronic Design, Cambridge, UK). The torque and rectified EMG data were then digitally filtered using a zero-phase infinite impulse response 4^th^-order Butterworth low-pass filter with a 4Hz corner frequency within MATLAB.

The processed baseline trials defined the torque offset and EMG threshold. Torque offset was defined as the average torque measured during the three baseline trials. EMG threshold, *EMG*_*t*_, was defined as:

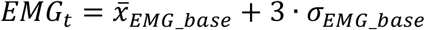

where 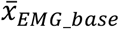 is the average of the three EMG baseline trials and 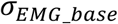 is the average EMG standard deviation over the three baseline trials.

For each static trial (i.e., the torque produced in a single combination of wrist and MCP joint positions), the processed torque and EMG data were divided into 1-second bins, resulting in 15 bins per trial. The average torque and EMG values were first calculated for each of the 15 bins. For a given 1-second bin in any trial, if the average EMG signal from any muscle exceeded *EMG*_*t*_ or if the torque deviated ≥ 5% from the mode across the entire trial, the bin was discarded. The average of the remaining bins within each trial were used to create the total torque versus wrist and MCP posture data set for each subject.

Differences in torques between limbs were calculated by subtracting non-paretic torque from paretic torque at each posture. Differences in MCP extension passive ROM (ePROM) between hands were calculated at each wrist posture. When data were missing or discarded, the difference was only calculated for postures where data was available in both paretic and non-paretic hands.

### Data Analysis

To test our primary hypothesis that passive torques in the paretic hand are greater than the non-paretic hand within each group (severe BoNT-injected, severe no-BoNT, moderate, and mild), we compared the differences between the participants’ paretic and non-paretic hands for MCP joint torques and wrist joint torques. Secondarily, we compared torque differences across groups and evaluated the interlimb differences for ePROM across all wrist postures. A distinct linear-mixed model (LMM) was implemented in SPSS (v26.0 IBM Corp Armonk, NY) for each parameter of interest (i.e., 3 models). LMMs allow for within-participant experimental designs by correcting for repeated measurements within participants while allowing for missing data points.^55^

When interlimb difference in MCP or wrist torques was the dependent variable, the linear-mixed model fixed effects included group, MCP position, wrist position, and their interactions. When interlimb difference in ePROM was the dependent variable, fixed effects included group, wrist position, and their interaction. An intercept random effect for participant was included in all models to account for random differences across participants.

Within each of the 4 impairment groups, post-hoc t-tests were used to test if the within-subject, interlimb torque differences were significantly different from zero. Significance was calculated using the T.DIST.RT function in Microsoft Excel (2020); inputs were the ratio of the marginal mean and standard error for each group and the degrees of freedom, all outputs from the LMM model in SPSS. To determine between group differences, post-hoc pairwise comparisons among the groups were performed. Significance of the pairwise comparisons were also calculated using the T.DIST.RT Excel function and outputs from the LMM models. To maintain p<0.05 as threshold of significance globally, the threshold for individual t-tests in the post-hoc analyses was reduced using the Bonferroni correction for multiple comparisions to p=0.005 (p=0.05/10). Across each dependent variable there were 10 post-hoc comparisons: the within-subject, different from zero comparison for each of the 4 groups, and 6 between group comparsions. Results are reported as average ± one standard deviation unless otherwise noted. Finally, to assess whether the differences in observed, mean, within-subject interlimb differences met our a priori assumptions, we calculated the Cohen’s d effect size achieved for each combination of wrist and MCP joint posture from the experimental data.

### Data availability

The data sets used within this analysis are available by request from the authors.

## RESULTS

### Passive Torque about the Wrist and MCP joints

Interlimb differences in MCP joint torques were small and were not significantly different for the mild (p= 0.188), moderate (p=0.046), or severe (p=0.017) impairment groups without a history of BoNT injections (cf., rows 1-3, Fig. 2a, p<0.005 is the threshold for significance for post-hoc t-tests). In contrast, the BoNT-injected group had significantly greater MCP joint torques in their paretic versus non-paretic hands (Fig. 2; p=5.83×10^−8^). The observed effects in interlimb differences for the the BoNT group were consistent with the large effect sizes (Cohen’s d = 1.1) we powered the study to detect. For example, the average of the achieved standardized effect size for all postures was 1.24±1.09 (range = 0.10-8.64), with effect sizes generally increasing with wrist and MCP extension. Effect sizes observed for interlimb MCP torque differences for the impairment groups without a history of BoNT were smaller, with mean Cohen’s d across all postures of 0.77±1.2, 0.49±0.33, and 0.47±0.63 for the severe, moderate, and mild groups, respectively. Interlimb differences in MCP torque increased with wrist and MCP extension for the BoNT group only (Fig 2b). The interlimb difference in MCP torques observed in the BoNT group were significantly greater than each no-BoNT group (p < 4.47×10^−4^ for all, Fig. 2b)

**Figure 2:**
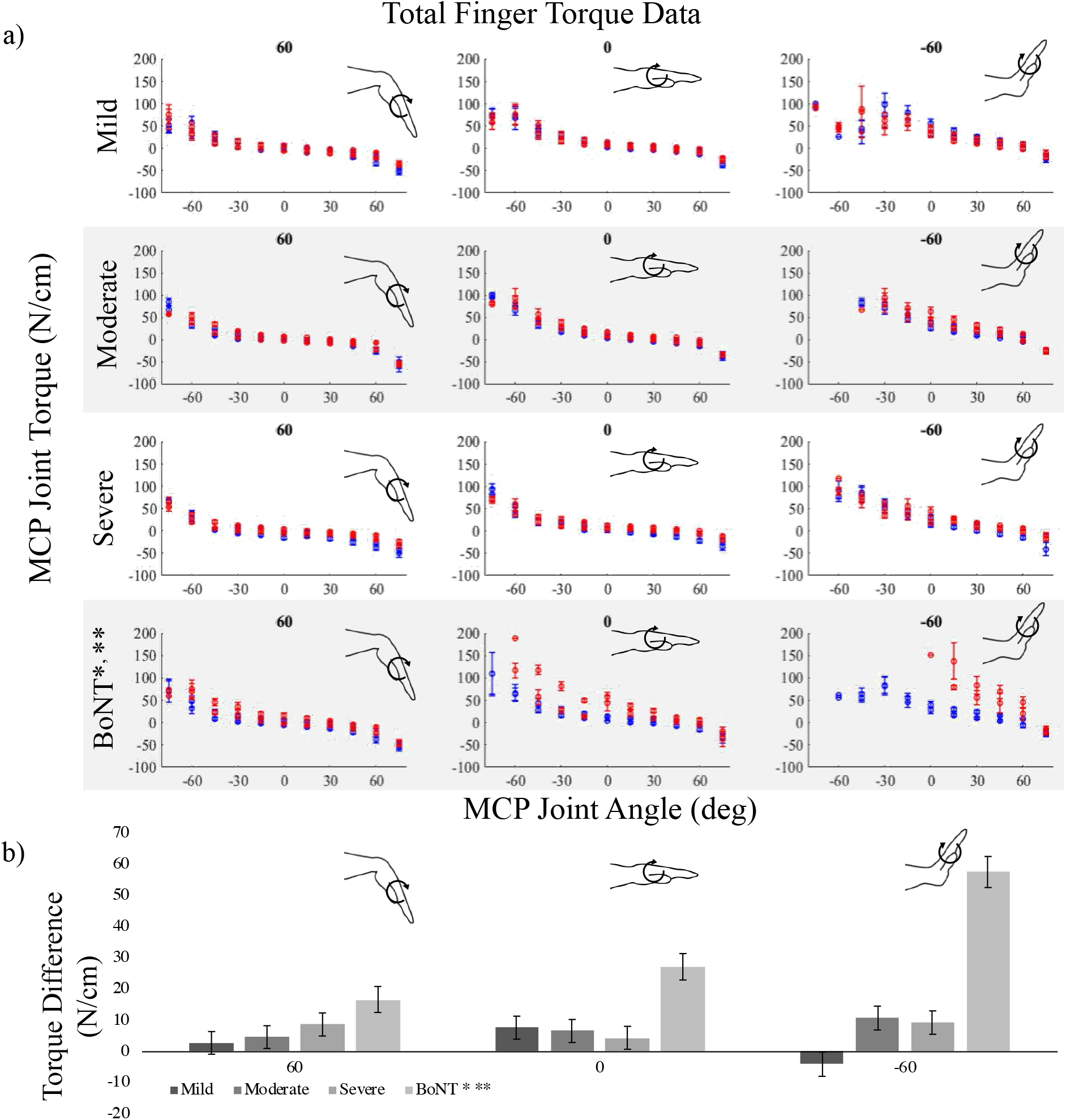
Average metacarpophalangeal (MCP) torques measured in the paretic (red) and non-paretic (blue) limbs over MCP joint range of motion for the (a) no-BoNT mildly, moderately, and severely impaired and BoNT-injected groups. Data are shown for 3 of the 9 wrist postures tested, including locked in 60 (left), 0 (middle), and -60 degrees flexion (right). Negative angles and torques indicate extension, positive indicates flexion. (b) Average interlimb difference in MCP torques (paretic torque minus non-paretic torque) for the same locked wrist postures in (a), for each group (shaded grey bars). * denotes significant difference (p<0.005) in torques measured between paretic and non-paretic limbs within the group. ** denotes significant difference (p<0.005) from all other groups. Only the BoNT-injected torque differences were significantly different between limbs and greater than the other groups. Error bars indicate one standard error of measurement.

Passive torques at the wrist replicated the findings at the MCP joints. Specifically, significant differences in passive wrist torques between limbs were only observed in the BoNT group (p = 4.51 x10^−6^; Fig. 3a). The distinction in standardized effect sizes observed in the BoNT group compared to the 3 no-BoNT groups was even greater for wrist torques than for MCP torques. The average, achieved standardized effect size (Cohen’s d) for interlimb wrist torque differences across all postures was 1.65±1.28 (range = 0.35-8.38) for the BoNT group compared to 0.30±0.35, 0.33±0.25 and 0.32±0.34 for the mild, moderate, and severe no-BoNT groups, respectively. Between groups, the interlimb difference in wrist torques observed in the BoNT group was significantly greater than each of the no-BoNT groups (p < 2.10×10^−3^ for all; Fig. 3b).

**Figure 3:**
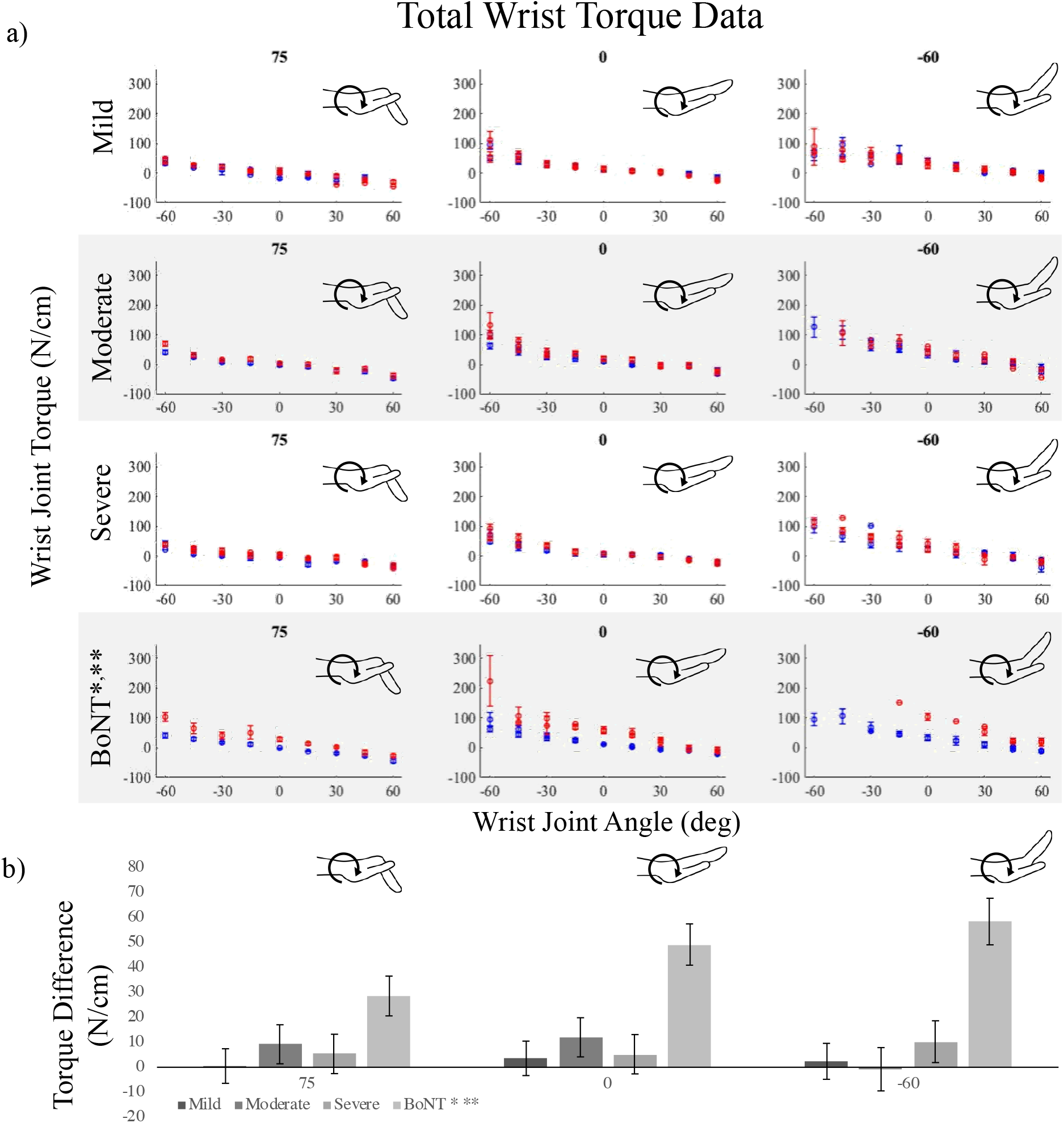
Average wrist torques measured in the paretic (red) and non-paretic (blue) limb over the wrist’s range of motion for the (a) no-BoNT mildly, moderately, and severely impaired and BoNT-injected groups. Metacarpophalangeal (MCP) posture was locked in -60 (left), 0 (middle), and 75 degrees flexion (right). Negative angles and torques indicate extension and positive flexion. (b) Average interlimb difference in wrist torques (paretic torques minus non-paretic torques) for each group (shaded grey bars), at each locked MCP posture. * denotes significant difference (p<0.005) in torques measured between paretic and non-paretic limbs within the group. ** denotes significant difference (p<0.005) from all other groups. Only the BoNT-injected torque differences were significantly different between limbs and greater than the other groups. Error bars indicate one standard error of measurement.

### Impact on Passive Range of motion

In general, the observed effects for passive MCP extension ROM in the three no-BoNT groups were small, especially when compared to the BoNT group. For the severe and mild no-BoNT groups, the passive limits of MCP extension were not significantly different between limbs; with ePROM differences of -3.15°±19.02° (p=0.216) and 2.47°±9.12° (p=0.274) and standardized effect sizes of 0.24±0.23 and 0.30±0.24 averaged across the 9 wrist postures tested, respectively. For the moderate no-BoNT group, the limit of passive MCP extension was 12.0°±3.9° less extended in the paretic limb than the non-paretic limb (p = 2.18×10^−3^), a significant interlimb difference that was on the same order as the measurement resolution for ePROM in this study (15°). In contrast, for the BoNT-injected group, the interlimb difference in ePROM across all wrist angles was much larger (−36.3°±4.5°; p=1.94×10^−9^).

## Discussion

The main objective of this study is to determine the extent to which the prolonged changes in neural input and muscle use experienced by individuals with a chronic hemiparetic stroke are associated with differences in muscle passive biomechanical properties between limbs. To this purpose, we evaluated a cohort of 27 individuals with chronic hemiparetic stroke, with no history of BoNT injections, spanning mild to severe hand impairments and an additional 7 individuals who had previously received BoNT injections, all with severe hand impairments. Our primary hypothesis was that passive torques in the paretic hand are greater than in the non-paretic hand. Strikingly, in the participants without BoNT, we did not observe significant interlimb passive torque differences at either the wrist or the MCP joints. Importantly, we powered the study to detect large effect sizes, and the significant results observed for the BoNT group demonstrate that we did enroll sufficient participant numbers to detect the large standardized effect sizes we expected to observe. However, the standardized effect sizes observed in the no-BoNT groups were much smaller than we anticipated. Overall, the small absolute magnitude of the interlimb torque differences (c.f. Figs 2b & 3b) paired with the small absolute magnitude of interlimb ePROM differences observed (c.f. Fig 4) among the no-BoNT groups suggest that the observed torque differences in the paretic limb are not a critical impediments to passive ROM after a stroke.

**Figure 4:**
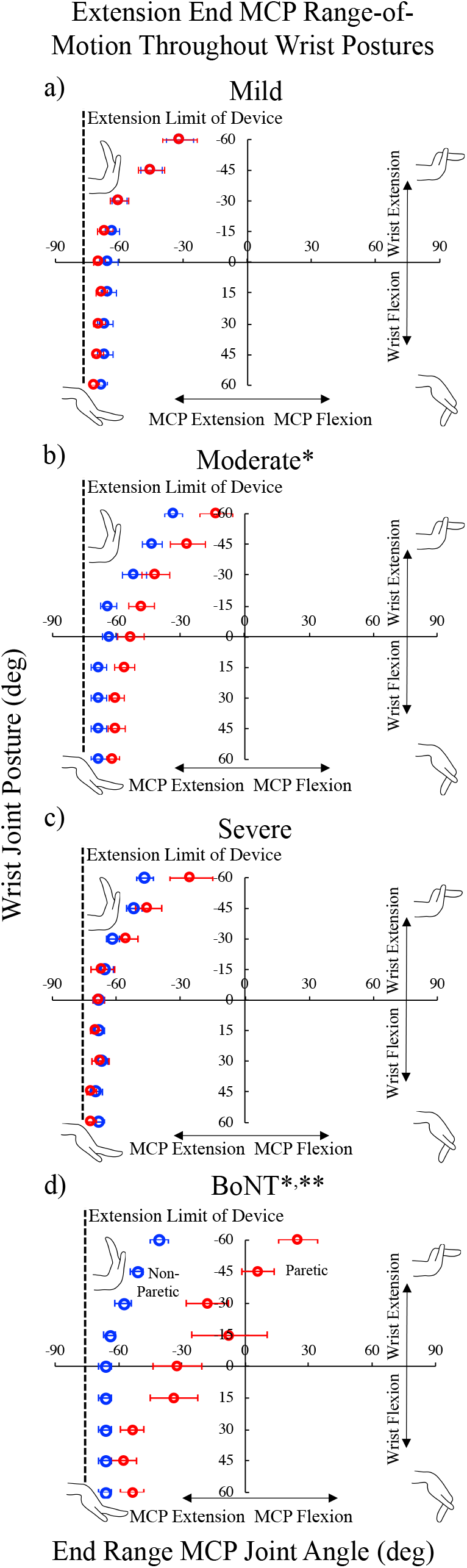
Limits of the metacarpophalangeal (MCP) passive range of motion (ROM) in extension throughout the custom device’s nine available wrist postures for the no-BoNT (a) mildly, (b) moderately, and (c) severely impaired and (d) BoNT-injected groups groups. All, but one BoNT-injected subject (limited to -15 degrees), could be positioned in all 9 wrist postures, limits of ROM for MCP presented are the average for the paretic (red) and non-paretic (blue) hands. MCP extension of the paretic hand in the BoNT-injected and moderate groups were significantly less than the non-paretic hand (*denotes p <0.005 between paretic v non-paretic hands). ** denotes significant difference (p<0.005) from all other groups. Deficits in passive extension increased with wrist extension. No significant loss of passive MCP extension was observed in the severe or mild groups. Error bars indicate one standard error of measurement.

Unlike many previous studies following chronic hemiparetic stroke,^7,30,31,33-37^ we did not observe greater passive torques in the paretic limb among the participants who had never received BoNT. Most of the previous studies evaluated lower extremity (ankle)^30,31,33,34,37^ or more proximal (elbow)^33,35^ joints; we studied distal wrist and finger joints. Following a stroke, the loss of corticospinal projections has a significantly greater impact on distal wrist and finger function than more proximal upper extremity joints.^56^ By replacing corticospinal drive with the indirect corticoreticulospinal system there is a significant neuromodulatory effect on spinal motorneurons due to greater release of monoamines (serotonin and norepinephrine) in the spinal cord.^57^ This increase of monoamines causes motoneuron hyperexcitability and increased involuntary muscle activation or hypertonia, which especially effects the distal wrist and finger muscles.^15,57,58^ This increased hypertonia and continuous activation may cause greater impairments of these more distal muscles during movement but it also may serve a protective effect on the muscle structure preventing significant atrophy and passive mechanical changes. This is an area that should be further explored and is currently under investigation within our lab.

Another factor contributing to the differences between our study and previous studies is the great care we took to reduce muscle hyperactivity during data collection. Inconsistencies between our results and previous studies may underscore the importance of this factor. Especially at the wrist and fingers, simple instructions to relax are likely insufficient. Many individuals with chronic stroke are unable to fully relax while awake due to reticulospinal mediated motoneuron hyperexcitability and muscle hypertonia. Decreasing hypertonia of muscles may require a significant reduction in corticoreticulospinal tract activity,^52-54^ that we addressed by encouraging participants to sleep during testing. A decrease in muscle activity was observed during test preparations (Figure 1), activity levels were strictly monitored throughout data collection. These critical steps should be considered when attempting to quantify passive muscle mechanics *in vivo* within any future study within a “spastic” population.

This study is also unique because we explicitly stratified participants based on prior clinical history of BoNT. Clinically, BoNT injections are frequently administered to alleviate muscle hyperactivity or hypertonia within the wrist and finger muscles. Approximately 45% of stroke survivors have spasticity^40,41^ and BoNT is the preferred treatment for decreasing muscle hyperactivity and increasing ROM. Each of our participants’ final BoNT injection was at least four years prior to our data collection (Table 2). Within our BoNT-injected group, we observed interlimb torque differences that were significantly larger than those observed in the no-BoNT groups. The BoNT-injected group was also the only group where we observed interlimb torque differences that systematically increased with wrist and MCP extension (Figures 2b & 3b). Extended postures lengthen the wrist and finger flexor muscles, which are the muscles that were injected with BoNT. Thus, our results suggest the possibility of a long lasting negative effect of BoNT: substantially increased passive muscle stiffness.

The increased passive joint torques we observed in the paretic limb in the BoNT group was also associated with substantial deficits in passive MCP extension relative to the nonparetic limb (Figure 4). This chronic limitation in ROM contrasts with acute increases in passive ROM, observed 1-2 months following BoNT injection.^59^ Importantly, the deficits in passive ROM we observed likely underestimate the extent that active ROM would be affected. We would expect even larger losses in active ROM as active finger extensor strength decreases by as much as 90%^6-8^ and therefore would likely be unable to overcome the increased passive torques about the fingers.

Previous animal studies demonstrate increased collagen content in the muscle following BoNT injections; these increases were observed within 6 months and lasted the animal’s lifespan.^22,23,26^ The underlying mechanism of this increased collagen content is unknown. Because BoNT chemically denervates a muscle the resulting adaptation may be similar to traditional muscle denervation. Denervated muscle has been shown to demonstrate increases in collagen content within a month after denervation^60,61^ and a recent study comparing muscle treated with BoNT and a denervated muscle demonstrated similar patterns of progressive atrophy and collagen content increases (Richard L. Lieber, personal communication of unpublished data, 2019). If BoNT is going to continue to be the ‘go to’ treatment for muscle hypertonia, further studies into the mechanism by which BoNT affects muscle are necessary to enhance our understanding of its long-term effects and to optimize recovery post stroke.

Because we quantified joint properties, we are unable to discern how mechanisms at the muscle level, such as atrophy (loss of contractile material), adaptations of muscle extra-cellular matrix (ECM), tendon compliance, or their combination) contribute to our results. In general, no study quantifying joint properties can distinguish between effects due to adaptations in muscle architecture, ECM, or tendon compliance. Future work could incorporate muscle imaging and tissue analyses to further quantify the impact of potential muscle volume and structural changes.

Another important limitation of this study is that it is a cross-sectional study. While we report striking differences between interlimb differences in passive joint torques and the range of motion between the BoNT group and the no-BoNT groups, our data does not establish causality between the BoNT injections and increased muscle or joint stiffness. Similarly, because this is not a prospective, longitudinal study we are not reporting observations of adaptations in chronic stroke muscle over time. Despite these limitations, the within subject control provided by the non-paretic limb provides a valuable comparison in our retrospective study.

Finally, based on our a priori expectations for interlimb differences, we powered the study to detect changes of large effect sizes. For the participants in our study without a previous history of BoNT, we generally observed effects that were smaller than our expectations. The data we present here could be leveraged to design future studies that would allow more robust statistical conclusions about such smaller effects. In general, despite this limitation, the smaller effects we observed lead us to conclude that the passive torque differences we observed in the paretic limb did not substantially limit passive ROM in these participants.

## Conclusions

To the best of our knowledge, the current study is the most thorough investigation of *in vivo* passive elastic torques at the hand in the chronic hemiparetic stroke population. Our findings indicate that, after stroke, prolonged altered use and neural inputs to muscle do not substantially increase or negatively impact the passive torques about either the wrist or fingers, nor significantly limit passive extension at the fingers, unless an individual has received BoNT. This suggests that clinically observed stiffness and loss of ROM is likely due to either neurally driven muscle hypertonia or long-term detrimental increases in muscle stiffness following BoNT injections. Additionally, this provides further evidence that in the absence of mechanical muscle alterations, the loss of hand function post-stroke is primarily due to weakness from voluntary activation deficits^6^ and impaired control of the muscles of the hand^11,56^ following disruptions of corticofugal motor pathways.^62,63^ Future rehabilitation techniques should therefore focus on motor deficits post stroke to achieve improved hand function rather than preventing passive mechanical changes. Additionally, the potential adverse effects of BoNT that our data highlights should be explored to determine if the positive effects of reduced muscle hypertonia and spasticity outweigh potential negative side effects of increased muscle stiffness in order to maximize recovery of individuals following a hemiparetic stroke.

## Data Availability

The data set used within this analysis is available by request from the authors.

## Acknowledgements

We especially want to thank Dr. Arno Stienen and Paul Krueger for their assistance with the custom device design and modification, and Vikram Darbhe for his assistance in experimental set-up and data collection

## References

1. Baker SN, Zaaimi B, Fisher KM, Edgley SA, Soteropoulos DS. Pathways mediating functional recovery. Prog Brain Res. 2015;218:389–412.

2. Zaaimi B, Edgley SA, Soteropoulos DS, Baker SN. Changes in descending motor pathway connectivity after corticospinal tract lesion in macaque monkey. Brain. 2012;135(Pt 7):2277–2289.

3. Ellis MD, Drogos J, Carmona C, Keller T, Dewald JP. Neck rotation modulates flexion synergy torques, indicating an ipsilateral reticulospinal source for impairment in stroke. J Neurophysiol. 2012;108(11):3096–3104.

4. McPherson JG, Chen A, Ellis MD, Yao J, Heckman CJ, Dewald JPA. Progressive recruitment of contralesional cortico-reticulospinal pathways drives motor impairment post stroke. J Physiol. 2018;596(7):1211–1225.

5. Karbasforoushan H, Cohen-Adad J, Dewald JPA. Brainstem and spinal cord MRI identifies altered sensorimotor pathways post-stroke. Nat Commun. 2019;10(1):3524.

6. Hoffmann G, Conrad MO, Qiu D, Kamper DG. Contributions of voluntary activation deficits to hand weakness after stroke. Top Stroke Rehabil. 2016;23(6):384–392.

7. Kamper DG, Fischer HC, Cruz EG, Rymer WZ. Weakness is the primary contributor to finger impairment in chronic stroke. Arch Phys Med Rehabil. 2006;87(9):1262–1269.

8. Kamper DG, Harvey RL, Suresh S, Rymer WZ. Relative contributions of neural mechanisms versus muscle mechanics in promoting finger extension deficits following stroke. Muscle Nerve. 2003;28(3):309–318.

9. Dewald JP, Pope PS, Given JD, Buchanan TS, Rymer WZ. Abnormal muscle coactivation patterns during isometric torque generation at the elbow and shoulder in hemiparetic subjects. Brain. 1995;118 (Pt 2):495–510.

10. Dewald JP, Sheshadri V, Dawson ML, Beer RF. Upper-limb discoordination in hemiparetic stroke: implications for neurorehabilitation. Top Stroke Rehabil. 2001;8(1):1–12.

11. Miller LC, Dewald JP. Involuntary paretic wrist/finger flexion forces and EMG increase with shoulder abduction load in individuals with chronic stroke. Clin Neurophysiol. 2012;123(6):1216–1225.

12. Sukal TM, Ellis MD, Dewald JP. Shoulder abduction-induced reductions in reaching work area following hemiparetic stroke: neuroscientific implications. Exp Brain Res. 2007;183(2):215–223.

13. Brunnstrom S. Movement therapy in hemiplegia: a neurophysiological approach. 1st ed. New York,: Medical Dept.; 1970.

14. Kamper DG, Rymer WZ. Quantitative features of the stretch response of extrinsic finger muscles in hemiparetic stroke. Muscle Nerve. 2000;23(6):954–961.

15. McPherson JG, Ellis MD, Heckman CJ, Dewald JP. Evidence for increased activation of persistent inward currents in individuals with chronic hemiparetic stroke. J Neurophysiol. 2008;100(6):3236–3243.

16. McPherson JG, Stienen AH, Drogos JM, Dewald JP. Modification of Spastic Stretch Reflexes at the Elbow by Flexion Synergy Expression in Individuals With Chronic Hemiparetic Stroke. Arch Phys Med Rehabil. 2017.

17. Bhadane MY, Gao F, Francisco GE, Zhou P, Li S. Correlation of Resting Elbow Angle with Spasticity in Chronic Stroke Survivors. Front Neurol. 2015;6:183.

18. O’Dwyer NJ, Ada L, Neilson PD. Spasticity and muscle contracture following stroke. Brain. 1996;119 (Pt 5):1737–1749.

19. Lee JM, Gracies JM, Park SB, Lee KH, Lee JY, Shin JH. Botulinum Toxin Injections and Electrical Stimulation for Spastic Paresis Improve Active Hand Function Following Stroke. Toxins (Basel). 2018;10(11).

20. Dong Y, Wu T, Hu X, Wang T. Efficacy and safety of botulinum toxin type A for upper limb spasticity after stroke or traumatic brain injury: a systematic review with meta-analysis and trial sequential analysis. Eur J Phys Rehabil Med. 2017;53(2):256–267.

21. Simpson DM, Hallett M, Ashman EJ, et al. Practice guideline update summary: Botulinum neurotoxin for the treatment of blepharospasm, cervical dystonia, adult spasticity, and headache: Report of the Guideline Development Subcommittee of the American Academy of Neurology. Neurology. 2016;86(19):1818–1826.

22. Minamoto VB, Suzuki KP, Bremner SN, Lieber RL, Ward SR. Dramatic changes in muscle contractile and structural properties after 2 botulinum toxin injections. Muscle Nerve. 2015;52(4):649–657.

23. Ward SR, Minamoto VB, Suzuki KP, Hulst JB, Bremner SN, Lieber RL. Recovery of rat muscle size but not function more than 1 year after a single botulinum toxin injection. Muscle Nerve. 2017.

24. Ates F, Yucesoy CA. Effects of botulinum toxin type A on non-injected bi-articular muscle include a narrower length range of force exertion and increased passive force. Muscle Nerve. 2014;49(6):866–878.

25. Yucesoy CA, Ateş F. BTX-A has notable effects contradicting some treatment aims in the rat triceps surae compartment, which are not confined to the muscles injected. J Biomech. 2018;66:78–85.

26. Thacker BE, Tomiya A, Hulst JB, et al. Passive mechanical properties and related proteins change with botulinum neurotoxin A injection of normal skeletal muscle. J Orthop Res. 2012;30(3):497–502.

27. Kaya CS, Yilmaz EO, Akdeniz-Dogan ZD, Yucesoy CA. Long-Term Effects With Potential Clinical Importance of Botulinum Toxin Type-A on Mechanics of Muscles Exposed. Front Bioeng Biotechnol. 2020;8:738.

28. Pingel J, Wienecke J, Lorentzen J, Nielsen JB. Botulinum toxin injection causes hyper-reflexia and increased muscle stiffness of the triceps surae muscle in the rat. J Neurophysiol. 2016;116(6):2615–2623.

29. Mathevon L, Michel F, Decavel P, Fernandez B, Parratte B, Calmels P. Muscle structure and stiffness assessment after botulinum toxin type A injection. A systematic review. Ann Phys Rehabil Med. 2015;58(6):343–350.

30. Gao F, Grant TH, Roth EJ, Zhang LQ. Changes in passive mechanical properties of the gastrocnemius muscle at the muscle fascicle and joint levels in stroke survivors. Arch Phys Med Rehabil. 2009;90(5):819–826.

31. Kwah LK, Herbert RD, Harvey LA, et al. Passive mechanical properties of gastrocnemius muscles of people with ankle contracture after stroke. Arch Phys Med Rehabil. 2012;93(7):1185–1190.

32. Nelson CM, Murray WM, Dewald JPA. Motor Impairment-Related Alterations in Biceps and Triceps Brachii Fascicle Lengths in Chronic Hemiparetic Stroke. Neurorehabil Neural Repair. 2018;32(9):799–809.

33. Given JD, Dewald JP, Rymer WZ. Joint dependent passive stiffness in paretic and contralateral limbs of spastic patients with hemiparetic stroke. J Neurol Neurosurg Psychiatry. 1995;59(3):271–279.

34. Mirbagheri MM, Alibiglou L, Thajchayapong M, Rymer WZ. Muscle and reflex changes with varying joint angle in hemiparetic stroke. J Neuroeng Rehabil. 2008;5:6.

35. Eby S, Zhao H, Song P, et al. Quantitative Evaluation of Passive Muscle Stiffness in Chronic Stroke. Am J Phys Med Rehabil. 2016;95(12):899–910.

36. de Gooijer-van de Groep KL, de Vlugt E, van der Krogt HJ, et al. Estimation of tissue stiffness, reflex activity, optimal muscle length and slack length in stroke patients using an electromyography driven antagonistic wrist model. Clin Biomech (Bristol, Avon). 2016;35:93–101.

37. Sinkjaer T, Magnussen I. Passive, intrinsic and reflex-mediated stiffness in the ankle extensors of hemiparetic patients. Brain. 1994;117 (Pt 2):355–363.

38. Freire B, Dias CP, Goulart NB, et al. Achilles tendon morphology, plantar flexors torque and passive ankle stiffness in spastic hemiparetic stroke survivors. Clin Biomech (Bristol, Avon). 2017;41:72–76.

39. Schmit BD, Dewald JP, Rymer WZ. Stretch reflex adaptation in elbow flexors during repeated passive movements in unilateral brain-injured patients. Arch Phys Med Rehabil. 2000;81(3):269–278.

40. Opheim A, Danielsson A, Alt Murphy M, Persson HC, Sunnerhagen KS. Upper-limb spasticity during the first year after stroke: stroke arm longitudinal study at the University of Gothenburg. Am J Phys Med Rehabil. 2014;93(10):884–896.

41. Wissel J, Manack A, Brainin M. Toward an epidemiology of poststroke spasticity. Neurology. 2013;80(3 Suppl 2):S13–19.

42. Levy J, Molteni F, Cannaviello G, Lansaman T, Roche N, Bensmail D. Does botulinum toxin treatment improve upper limb active function? Ann Phys Rehabil Med. 2019;62(4):234–240.

43. Hara T, Momosaki R, Niimi M, Yamada N, Hara H, Abo M. Botulinum Toxin Therapy Combined with Rehabilitation for Stroke: A Systematic Review of Effect on Motor Function. Toxins (Basel). 2019;11(12).

44. Lawrence ES, Coshall C, Dundas R, et al. Estimates of the prevalence of acute stroke impairments and disability in a multiethnic population. Stroke. 2001;32(6):1279–1284.

45. Broeks JG, Lankhorst GJ, Rumping K, Prevo AJ. The long-term outcome of arm function after stroke: results of a follow-up study. Disabil Rehabil. 1999;21(8):357–364.

46. Parker VM, Wade DT, Langton Hewer R. Loss of arm function after stroke: measurement, frequency, and recovery. Int Rehabil Med. 1986;8(2):69–73.

47. Nakayama H, Jorgensen HS, Raaschou HO, Olsen TS. Recovery of upper extremity function in stroke patients: the Copenhagen Stroke Study. Arch Phys Med Rehabil. 1994;75(4):394–398.

48. Gowland C, Stratford P, Ward M, et al. Measuring Physical Impairment and Disability with the Chedoke-Mcmaster Stroke Assessment. Stroke. 1993;24(1):58–63.

49. Knutson JS, Kilgore KL, Mansour JM, Crago PE. Intrinsic and extrinsic contributions to the passive moment at the metacarpophalangeal joint. J Biomech. 2000;33(12):1675–1681.

50. Faul F, Erdfelder E, Buchner A, Lang AG. Statistical power analyses using G*Power 3.1: tests for correlation and regression analyses. Behav Res Methods. 2009;41(4):1149–1160.

51. Stienen A, Moulton TS, Miller LC, Dewald JPA. Wrist and Finger Torque Sensor for the Quantification of Upper Limb Motor Impairments Following Brain Injury. Paper presented at: 2011 IEEE International Conference on Rehabilitation Robotics2011; Zurich, Switzerland.

52. Fraigne JJ, Torontali ZA, Snow MB, Peever JH. REM Sleep at its Core - Circuits, Neurotransmitters, and Pathophysiology. Front Neurol. 2015;6:123.

53. Krenzer M, Anaclet C, Vetrivelan R, et al. Brainstem and spinal cord circuitry regulating REM sleep and muscle atonia. PLoS One. 2011;6(10):e24998.

54. Hodes R, Dement WC. Depression of Electrically Induced Reflexes (“H-Reflexes”) in Man during Low Voltage Eeg “Sleep”. Electroencephalogr Clin Neurophysiol. 1964;17:617–629.

55. Magezi DA. Linear mixed-effects models for within-participant psychology experiments: an introductory tutorial and free, graphical user interface (LMMgui). Front Psychol. 2015;6:2.

56. Lan Y, Yao J, Dewald JPA. The Impact of Shoulder Abduction Loading on Volitional Hand Opening and Grasping in Chronic Hemiparetic Stroke. Neurorehabil Neural Repair. 2017;31(6):521–529.

57. Li S, Chen YT, Francisco GE, Zhou P, Rymer WZ. A Unifying Pathophysiological Account for Post-stroke Spasticity and Disordered Motor Control. Front Neurol. 2019;10:468.

58. McPherson LM, Dewald JPA. Differences between flexion and extension synergy-driven coupling at the elbow, wrist, and fingers of individuals with chronic hemiparetic stroke. Clin Neurophysiol. 2019;130(4):454–468.

59. Hallett M. Explanation of timing of botulinum neurotoxin effects, onset and duration, and clinical ways of influencing them. Toxicon. 2015;107(Pt A):64–67.

60. Liu F, Tang W, Chen D, et al. Expression of TGF-beta1 and CTGF Is Associated with Fibrosis of Denervated Sternocleidomastoid Muscles in Mice. Tohoku J Exp Med. 2016;238(1):49–56.

61. Faturi FM, Franco RC, Gigo-Benato D, et al. Intermittent stretching induces fibrosis in denervated rat muscle. Muscle Nerve. 2016;53(1):118–126.

62. Kuypers HG. The Descending Pathways to the Spinal Cord, Their Anatomy and Function. Prog Brain Res. 1964;11:178–202.

63. Lawrence DG, Kuypers HG. The functional organization of the motor system in the monkey. I. The effects of bilateral pyramidal lesions. Brain. 1968;91(1):1–14.

